# Genomic epidemiology identifies emergence and rapid transmission of SARS-CoV-2 B.1.1.7 in the United States

**DOI:** 10.1101/2021.02.06.21251159

**Authors:** Nicole L. Washington, Karthik Gangavarapu, Mark Zeller, Alexandre Bolze, Elizabeth T. Cirulli, Kelly M. Schiabor Barrett, Brendan B. Larsen, Catelyn Anderson, Simon White, Tyler Cassens, Sharoni Jacobs, Geraint Levan, Jason Nguyen, Jimmy M. Ramirez, Charlotte Rivera-Garcia, Efren Sandoval, Xueqing Wang, David Wong, Emily Spencer, Refugio Robles-Sikisaka, Ezra Kurzban, Laura D. Hughes, Xianding Deng, Candace Wang, Venice Servellita, Holly Valentine, Peter De Hoff, Phoebe Seaver, Shashank Sathe, Kimberly Gietzen, Brad Sickler, Jay Antico, Kelly Hoon, Jingtao Liu, Aaron Harding, Omid Bakhtar, Tracy Basler, Brett Austin, Magnus Isaksson, Phillip G. Febbo, David Becker, Marc Laurent, Eric McDonald, Gene W. Yeo, Rob Knight, Louise C. Laurent, Eileen de Feo, Michael Worobey, Charles Chiu, Marc A. Suchard, James T. Lu, William Lee, Kristian G. Andersen

**Affiliations:** Helix, San Mateo, CA; Department of Immunology and Microbiology, The Scripps Research Institute, La Jolla, CA; Department of Ecology and Evolutionary Biology, University of Arizona, Tucson, AZ; Department of Laboratory Medicine, University of California San Francisco, San Francisco, CA; University of California, San Diego, CA; Illumina, San Diego, CA; Sharp Healthcare, San Diego, CA; San Diego County Health and Human Services Agency, San Diego, CA; Innovative Genomics Institute, Berkeley, CA; Department of Biostatistics, Fielding School of Public Health, and Departments of Biomathematics and Human Genetics, David Geffen School of Medicine, University of California, Los Angeles, Los Angeles, CA; Scripps Research Translational Institute, La Jolla, CA; Department of Integrative, Structural and Computational Biology, The Scripps Research Institute, La Jolla, CA 92037, USA

**Author notes:** Co-first authors (Nicole L. Washington, Karthik Gangavarapu). Co-senior authors (William Lee, Kristian G. Andersen).

## Abstract

As of January of 2021, the highly transmissible B.1.1.7 variant of SARS-CoV-2, which was first identified in the United Kingdom (U.K.), has gained a strong foothold across the world. Because of the sudden and rapid rise of B.1.1.7, we investigated the prevalence and growth dynamics of this variant in the United States (U.S.), tracking it back to its early emergence and onward local transmission. We found that the RT-qPCR testing anomaly of S gene target failure (SGTF), first observed in the U.K., was a reliable proxy for B.1.1.7 detection. We sequenced 212 B.1.1.7 SARS-CoV-2 genomes collected from testing facilities in the U.S. from December 2020 to January 2021. We found that while the fraction of B.1.1.7 among SGTF samples varied by state, detection of the variant increased at a logistic rate similar to those observed elsewhere, with a doubling rate of a little over a week and an increased transmission rate of 35-45%. By performing time-aware Bayesian phylodynamic analyses, we revealed several independent introductions of B.1.1.7 into the U.S. as early as late November 2020, with onward community transmission enabling the variant to spread to at least 30 states as of January 2021. Our study shows that the U.S. is on a similar trajectory as other countries where B.1.1.7 rapidly became the dominant SARS-CoV-2 variant, requiring immediate and decisive action to minimize COVID-19 morbidity and mortality.

## Introduction

Since the onset of the COVID-19 pandemic, there has been concern about novel SARS-CoV-2 variants emerging that are more transmissible. During the third quarter of 2020, the SARS-CoV-2 Variant of Concern (VOC) 202012/01 (a.k.a. 501Y.V1; B.1.1.7) lineage carrying the N501Y mutation emerged and took hold in the UK, followed by several European countries. The N501Y mutation is also shared with other VOCs first identified in South Africa (501Y.V2; B.1.351) (Tegally et al., 2020) and Brazil (501Y.V3; P.1) (Faria et al., 2021), but B.1.1.7 has several additional ‘signature’ mutations in the SARS-CoV-2 Spike protein, including deletions at 69-70 and 144, as well as mutations A570D, D614G, P681H, T716I, S982A, and D1118H (Rambaut et al., 2020a). The earliest sequence of B.1.1.7 was collected on September 20, 2020 in England (GISAID EPI_ISL_601443), but it has since spread rapidly across the UK, becoming the dominant lineage within just a few months (Chand et al., 2020; Cyranoski, 2021; ECDC, 2021; Rambaut et al., 2020a). This lineage is more transmissible, with a growth rate that has been estimated to be 40-70% higher than other SARS-CoV-2 lineages in multiple countries, which is hypothesized to be partly due to the N501Y mutation increasing receptor binding affinity of the SARS-CoV-2 spike protein with ACE2 (Volz et al., 2021). While initially thought to have comparable clinical outcomes to other SARS-CoV-2 variants, preliminary reports indicate that infection with B.1.1.7 may lead to ∼30% higher mortality rates (Iacobucci, 2021). Though the exact origin of the B.1.1.7 variant is unclear, it was only through the proactive and ongoing large scale SARS-CoV-2 genomic surveillance program in the U.K. facilitated the initial detection after investigators in South Africa had observed an association between N501Y and increased transmission (Chand et al., 2020).

Routinely administered RT-qPCR SARS-CoV-2 diagnostic tests can provide hints to the presence of viral lineages with sequence-based differences when mutations occur at the test’s target probe locations. Importantly, the 69-70 deletion, present in B.1.1.7 and other variants, can be characterized by the failure to detect the S gene in these tests, known as S gene target failure (SGTF) (Bal et al., 2020). Retrospective analyses from the U.K. show that the proportion of B.1.1.7 in SGTF samples rose from 3% during the week of October 12, 2020 to more than 90% during the week of November 30, 2020. It has since reached near-fixation across most of the U.K. (Chand et al., 2021).

Because of the sudden and rapid rise of the B.1.1.7 variant across the world, we sought to understand the prevalence and growth dynamics of this variant in the U.S., from early emergence to rapid onward transmission. Surveillance programs typically select a subset of RT-PCR SARS-CoV-2 tests for sequencing, and therefore identifying SGTF samples for sequencing provides a built-in prioritization method to enrich for detection of B.1.1.7. In this study we describe the introduction and early spread of B.1.1.7 in the U.S., based on historical SGTF rates in RT-PCR SARS-CoV-2 tests and a nationwide SGTF viral sequencing program. We find that B.1.1.7 arrived in the U.S. towards the end of November, 2020 and as of January, 2021 it has since spread to at least 30 U.S. states. Importantly, we find that the B.1.1.7 variant is 35-45% more transmissible across the country, doubling in relative frequency about every week and a half. These findings show that B.1.1.7 will likely become the dominant variant in many U.S. states by March, 2021, leading to further surges of COVID-19 in the country, unless urgent mitigation efforts are immediately implemented.

## Results

### The proportion of SGTF samples is rapidly increasing in the U.S

We examined the prevalence of SGTF in all SARS-CoV-2 positive samples from across the U.S. tested at Helix facilities since July 2020 (∼0.5 million samples; **Figure 1A**). The Helix^®^ COVID-19 Test calls positive samples when at least two of three targets (N, Orf1ab, and S) are detected using the Thermo Fisher TaqPath™ assay. We only considered samples to be SGTF if they were positive for both N and Orf1ab, and negative for S. We restricted our analyses to positive samples with Cq<27 for the N gene based on previous reports that single target failures were more frequent at higher Cq (Bal et al., 2020; Kara Steel And, 2021). We began to observe consistent, low frequency SGTF in early October 2020, with 0.2% of daily SARS-CoV-2-positive tests exhibiting this pattern during the week of October 18, 2020, followed by a steady increase (**Supplemental Figure S1**). We found that during the month of January 2021, the nationwide proportion of SGTF increased from an average of 0.8% in the first week to 4.2% in the last week (**Supplemental Figure S1**). As described in more detail below, the proportion of B.1.1.7 cases reached 3.6% in the U.S. by the last week of January, 2021 (**Figure 1B, C**).

**Figure 1.**
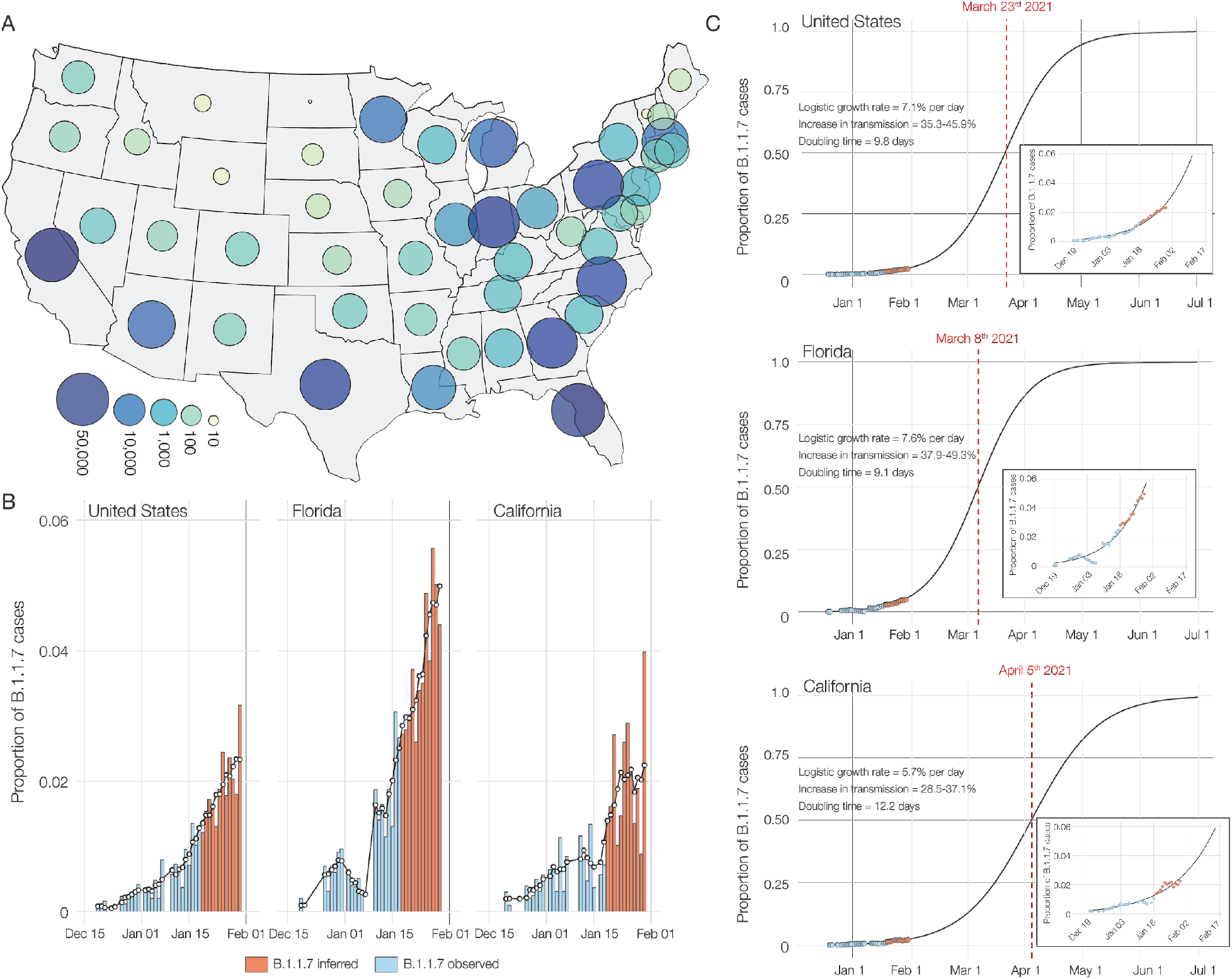
SGTF and B.1.1.7 in SARS-CoV-2 tests at Helix since December 15, 2020. **(A)** Map of contiguous states in the USA with each bubble representing the number of positive tests from each state. **(B)** Estimated proportion of B.1.1.7 in total number of positive tests with Cq(N gene) < 27, in the U.S., California and Florida from December 15th, 2020 to January 30th, 2021. The proportion of B.1.1.7 samples was estimated using: (Observed B.1.1.7 sequences/Sequenced SGTF samples) * (Positive tests with SGTF/Total positive tests). Due to the lag in sequencing, the average proportion of B.1.1.7 sequences in sequenced samples with SGTF from the last five days (January 13-18) was used to infer the proportion of B.1.1.7 cases in total positive tests for the January 19-30 time period between. The black line shows the 5-day rolling average of the estimated proportion of B.1.1.7 in total positives. **(C)** Logistic growth curves fit to the rolling average of the estimated proportion of B.1.1.7 in total positives for the U.S., Florida and California. The predicted time when the estimated proportion of B.1.1.7 cases crosses 0.5 is indicated in red.

To investigate regional differences across the U.S., we examined the nationwide distribution of SGTF. By grouping our samples based on patient state of residence, we observed SGTF in 22 out of 53 U.S. States and territories during January 2021, several of which had SGTF frequencies consistently above 1% (**Supplemental Figure S1**). When restricting our analysis to assess U.S. states with more than 500 positive tests in January 2021, we observed that the fraction of SGTF varies significantly across the nation (**Supplemental Figure S1**). A caveat to this analysis is that our testing footprint does not evenly cover the U.S. (**Figure 1A**), hence SGTF may currently be undetected in several states.

### Identification of B.1.1.7 using SARS-CoV-2 sequencing

Since SGTF only tracks the presence of the 69-70 deletion in the SARS-CoV-2 spike gene, it is not specific to B.1.1.7 (Washington et al., 2020). To investigate the proportion of B.1.1.7 in our SGTF samples, we sequenced all SGTF samples from December, 2020 through January, 2021.

We found that of the 460 SGTF samples we successfully sequenced, 209 (45%) were from the B.1.1.7 variant (insert; **Figure 2A** and **Supplemental Table S1**), distributed across ten U.S. states (insert; **Figure 2A**). In addition to the SGTF samples, we also sequenced three B.1.1.7 genomes that were detected as part of random SARS-CoV-2 genomic surveillance in California (**Supplemental Table S1**). Of the 212 B.1.1.7 sequences, 96 came from California and 87 Florida (insert; **Figure 2A**).

**Figure 2.**
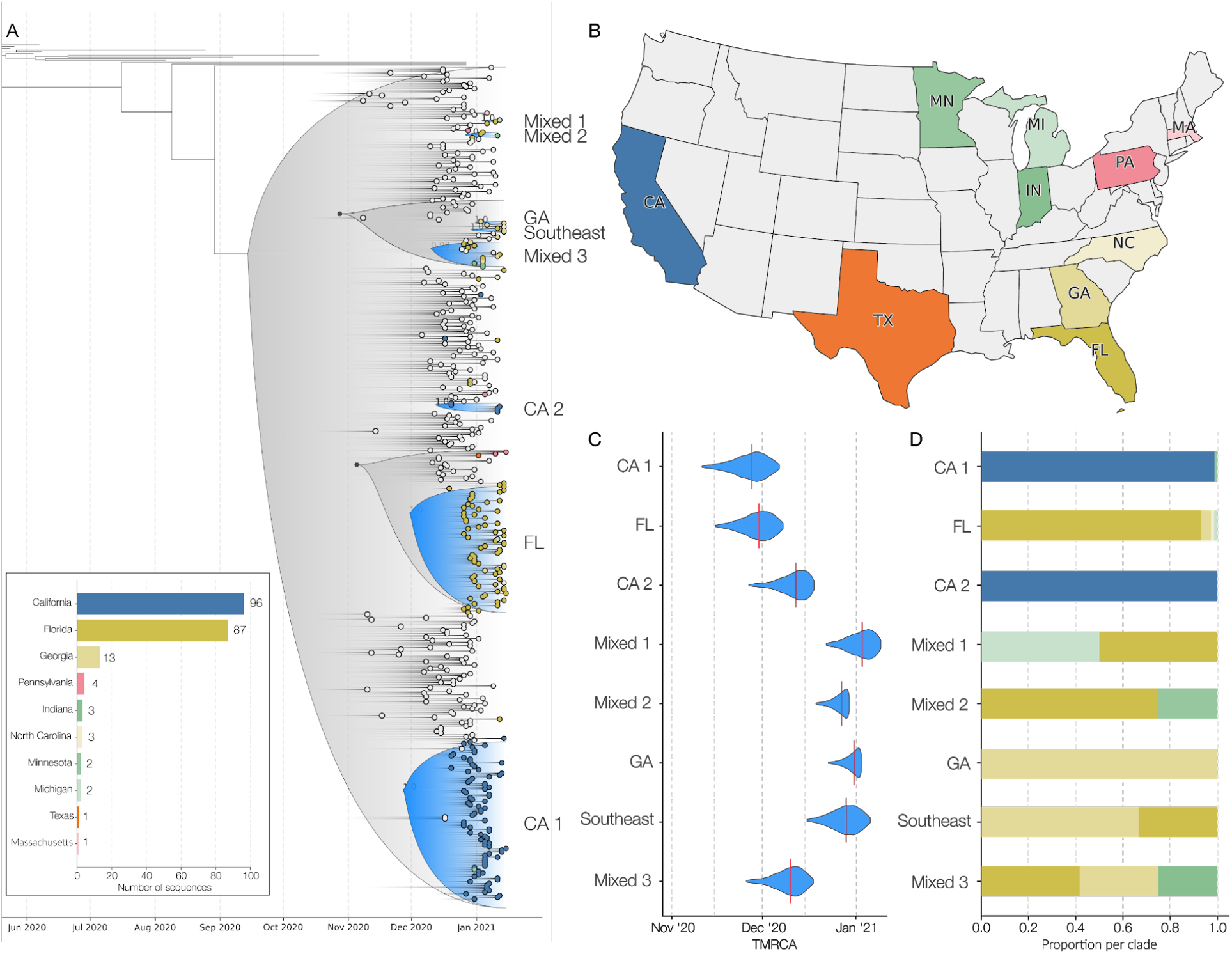
Phylogenetic analysis of B.1.1.7 lineage in the USA. (**A**) Maximum clade credibility (MCC) tree of the time resolved phylogenetic analysis of B.1.1.7 sequences in the U.S. in the context of sequences sampled globally. The gradient represents uncertainty in the tree topology. Clades that consist primarily of sequences sampled in the U.S. supported by a basal node with posterior probability ≥0.98 are colored in blue. The closest ancestral node to each clade with a posterior probability ≥ 0.98 is highlighted in black. (**B**) The color scheme of terminal nodes sampled in the MCC tree. Sequences sampled outside the U.S. are colored in light gray. States with no B.1.1.7 sequence sampling in the dataset are shown in white. (**C**) The TMRCA of each clade highlighted in the MCC tree. (**D**) The proportion of the geographic sampling of sequences within each clade (singletons have been excluded, including those in Texas, Pennsylvania, and Massachusetts). The colors follow the same scheme as shown in panel B.

We found that all the SARS-CoV-2 variant genomes from the U.S. contained all the B.1.1.7 ‘signature’ mutations, including del69-70, del144, N501Y, A570D, D614G, P681H, T716I, S982A, and D1118H (Rambaut et al., 2020a). None of our B.1.1.7 sequences contained any of the key mutations identified in the B.1.351 and P.1 variants, such as L18F, K417N/T, or E484K (Faria et al., 2021; Tegally et al., 2020), with the latter recently having been identified in B.1.1.7 lineages in the U.K. (Public Health England, 2020). We did, however, observe an additional nonsynonymous mutation, K1191N, in the majority of B.1.1.7 genomes from Florida, and one instance of Q493K, which has been implicated in immune escape (Starr et al., 2021).

In addition to the ten states with evidence of B.1.1.7 transmission identified in this study, we note that additional B.1.1.7 sequences have been reported from other testing labs, with 33 of 53 U.S. states and territories reporting to the CDC at least one case to date (CDC, 2021).

#### B.1.1.7 has an increased growth rate in the U.S. compared to non-B.1.1.7 lineages

To investigate the dynamics of B.1.1.7 in the U.S., we estimated the proportion of the variant out of total positive tests by multiplying the proportion of B.1.1.7 in all sequenced SGTF cases by the proportion of SGTF in all positive tests and taking the 5-day rolling average until January 18th. Given the lag of sequencing, we used the average proportion of B.1.1.7 in all sequenced SGTF cases from January 13 to January 18 to estimate the proportion of the variant out of total tests from January 18 to January 20. We found that the proportion of B.1.1.7 in our SGTF samples grew to a nationwide average of ∼90% in the middle of January, 2021, although with substantial variance across the country, ranging from ∼95% in California to ∼70% in Florida (**Supplemental Figure S2**).

Following the same methodology to estimate the proportion of B.1.1.7 in the population, we found that by the last week of January 2021, B.1.1.7 made up an average of ∼2.1% COVID-19 cases in the U.S., with ∼2.0% of all cases in California and ∼4.5% of all cases in Florida caused by B.1.1.7 (**Figure 1B**). To investigate the growth rate of B.1.1.7 in the U.S. compared to non-B.1.1.7 lineages, we fitted a logistic growth model to our rolling 5-day average of estimated B.1.1.7 cases. We determined that B.1.1.7 has a logistic growth rate of 0.07 in the U.S. (**Figure 1C**), which translates to an increased transmissibility of 35-46% in the U.S., with rates in California of 29-37% and 38-49% in Florida (**Figure 1C**). Using a serial interval of 5 - 6.5 days (Volz et al., 2021), we estimated the doubling time of B.1.1.7 to be 9.8 days in the U.S., 12.2 days in California, and 9.1 days in Florida (**Figure 1C**).

#### B.1.1.7 was introduced into the U.S. multiple times in November, 2020

To investigate the timing and number of introductions that led to the emergence of B.1.1.7 in the U.S., we combined our 212 B.1.1.7 genomes with a representative sampling of 292 additional sequences from outside the U.S. We used this dataset to reconstruct a Bayesian time-resolved phylogeny using BEAST (Suchard et al., 2018). We found that the majority of B.1.1.7 sequences from the U.S. cluster into two main clades (**Figure 2A**) with independent introductions into California (CA1; **Figure 2A**) and Florida (FL; **Figure 2A**).

In addition to these major clades, we also identified six smaller clades representing at least eight independent introductions (**Figure 2A**), as well as nineteen singletons showing additional introductions (**Figure 2A**) across the U.S. states we surveyed (**Figure 2B**). The “GA” clade (**Figure 2A**) contains three sequences and represents an independent introduction into Georgia. Clade “Southeast” (**Figure 2A**) contains two sequences from Georgia and one from Florida and represents an independent introduction into the Southeastern U.S. The “CA2” clade was made up of two sequences from California and constituted a separate introduction (**Figure 2A**), although these samples came from individuals with recent travel history to the U.K.

We found that the earliest timing of introductions into the U.S. was represented by the “CA1” clade, which had a median time to the most recent common ancestor (TMRCA; which depicts the likely start of sustained local transmission in California (Grubaugh et al., 2019a)) of November 27, 2020 (95% highest posterior probability (HPD): [November 14 - December 8]), followed by “FL” with a median TMRCA of November 29, 2020 (95% HPD: [November 17 - December 8]) (**Figure 2C**). Clade “GA” had a median TMRCA of December 31, 2020 (95% HPD: [December 22 - January 2]). We found that the other U.S. clades had median TMRCAs in December, 2020 and January, 2021, suggesting repeated introductions of B.1.1.7 into the U.S. from international locations from November, 2020 through present time.

#### B.1.1.7 has likely been spreading between U.S. states since late 2020

In addition to the main B.1.1.7 clades that contained sequences primarily from individual states, including “CA1”, “CA2”, “FL”, and “GA”, we found that clades “Mixed-1”, “Mixed-2”, “Mixed-3”, and “Southeast” were diverse with respect to geographic sampling, containing SARS-CoV-2 genomes from across multiple U.S. states (**Figure 2A**). These findings indicate that B.1.1.7 has been spreading locally between different U.S. states (**Figure 2D**), likely since at least December, 2020 based on our TMRCA estimates (**Figure 2A**).

Undersampling and bias in our B.1.1.7 sequencing make it unfeasible to estimate directionality and connectedness across the U.S., but the clustering of B.1.1.7 sequences into mixed clades suggest movement of the virus between Florida and Georgia, Florida and Minnesota, Florida and Michigan, and between the Southeastern states of Florida, Georgia, and North Carolina (**Figure 2D**).

## Discussion

The B.1.1.7 variant established itself as the dominant SARS-CoV-2 lineage in the U.K. within a couple of months after its detection (Bal et al., 2020; Chand et al., 2020). Since then, the variant has been increasingly observed across many European countries, including Portugal and Ireland, which, like the U.K., observed devastating waves of COVID-19 after B.1.1.7 became dominant (ECDC, 2021). In this study, we show that B.1.1.7 is currently at a relatively low frequency in the U.S., but our estimates show that its growth rate is at least 35-45% increased and doubling every week and a half. These findings are consistent with those from other countries (Volz et al., 2021), and given the current trajectory of B.1.1.7 in the U.S., it is almost certainly destined to become the dominant SARS-CoV-2 lineage by March, 2021 across many U.S. states, which is consistent with modeling analyses from the U.S. Centers for Disease Control and Prevention (Galloway et al., 2021). However, obtaining accurate estimates of the prevalence of B.1.1.7 across the country is complicated by uneven and biased sampling, combined with a lack of a national SARS-CoV-2 genomics surveillance program, like those implemented in the U.K., Denmark, Australia, New Zealand, among other places. In the current study, we obtain relatively robust estimates from California and Florida, but our analyses outside these places are limited.

Our phylogenetic analyses indicate that there have been multiple introductions of B.1.1.7 into the U.S., with the earliest dating back to the end of November, 2020. These analyses revealed large clades of closely related SARS-CoV-2 lineages clustering within individual states, as well as national spread indicated by several smaller clades defined by mixtures of samples from patients who reside in different U.S. states. These findings are consistent with community transmission following several of these introductions, including spread across U.S. states However, unresolved polytomies of sequences belonging to different regions nationally and internationally at several basal nodes in our phylogenies, mean that we are unable to fully resolve directionality and likely origins for the U.S. sequences (Grubaugh et al., 2019a). However, our TMRCA estimates coincide with increased periods of travel, where the U.S. Transportation Security Administration reported over one million travelers crossing checkpoints for several days during the peak Thanksgiving season (November 20-29, 2020) and for twelve of eighteen days surrounding the Christmas and New Year’s holidays (December 18, 2020 to January 4, 2021) (TSA, 2021), providing a likely explanation for how B.1.1.7 may have been introduced via international travel and spread across the U.S. via domestic travel.

In addition to well-supported local clades in California, Florida, and Georgia from our phylogenetic analyses, many of the initial B.1.1.7 cases in the U.S. did not report recent international travel prior to infection (Davis, 2020; Romo, 2020). These findings suggest that significant community transmission of the B.1.1.7 variant is already ongoing across the U.S., which is likely fueled by its increased growth rate and transmissibility of B.1.1.7. We found the growth rates in California (∼6% / day) to be slightly lower than those in Florida (∼8% / day). This difference may be due to differences in statewide or regional social distancing protocols or mobility patterns, population density, biases in sampling and/or demographics, or competition from other SARS-CoV-2 variants, like the B.1.429 variant recently described in California (CDPH, 2021). The nationwide growth rate of ∼7% / day is a little lower than those observed in Portugal (10% / day (Borges, 2021)), Denmark (10.3% / day (Statens Serums Institute)) and the United Kingdom (10.4% / day (Davies et al., 2020)). The reason for the slightly lower growth rate of B.1.1.7 in the U.S. compared to European countries require further investigation, but may be down to the relative sparsity of the current dataset, the lack of strong mitigation efforts in this country, or competition from other more transmissible SARS-CoV-2 variants.

Our study shows that although SGTF is not yet a universal proxy for the B.1.1.7 variant in the U.S., the increased growth rate of B.1.1.7 compared to other SGTF variants (Volz et al., 2021) will likely lead to B.1.1.7 overtaking other SGTF variants in a matter of weeks. This will allow the simple SGTF testing anomaly to be used to monitor the spread of B.1.1.7 in the U.S. in the absence of nationwide genomic surveillance.

While B.1.1.7 is of current interest given the concerns regarding increased transmission dynamics, the results here reinforce the need for ongoing SARS-CoV-2 genomic surveillance to monitor the dynamics of B.1.1.7 and other emerging SARS-CoV-2 variants, including those yet to be discovered. Because laboratories in the U.S. are only sequencing a small subset of SARS-CoV-2 samples, the true sequence diversity of SARS-CoV-2 in this country is still unknown. The more established surveillance programs in other countries have provided important warnings about variants of concern that can impact the U.S., with B.1.1.7 representing only one variant that demonstrates the capacity for exponential growth. As viral surveillance efforts increase in the U.S., we will no doubt find additional SARS-CoV-2 variants, some at high prevalence and others with increased fitness. Only with consistent, unbiased sequencing at scale that includes all geographic and demographic populations including those often underrepresented, together with continued international scientific collaborations and open data sharing, will we be able to accurately assess and follow new variants that emerge during the COVID-19 pandemic. Given that the SARS-CoV-2 VOCs B.1.1.7, B.1.351, and P.1 are still at relatively low frequency in the U.S., there is still time to implement the necessary surveillance programs and mitigation efforts in the weeks to come. Unless decisive and immediate public health action is taken, the increased transmission rate of these lineages and resultant higher effective reproduction number of SARS-CoV-2 will likely have devastating consequences to COVID-19 mortality and morbidity in the U.S. in a few months, if decisive action is not immediately taken.

## Supporting information

Supplemental Figure S1 - Percentage of SGTF for states with >500 positive tests

Supplemental Figure S2 - Percentage of B.1.1.7 in sequenced samples with SGTF over time

Supplemental Table S2 - GISAID Acknowledgements

Supplemental Table S1 - Sequence stats

## Data Availability

Raw data and processing scripts are available at https://github.com/andersen-lab/paper_2021_early-b117-usa;
Viral sequences have been deposited at GISAID, with accession numbers as listed in Supplemental Tables.;
Additionally, continuing monitoring of the phenomena described in the manuscript can be found at https://www.helix.com/covid19db

https://www.helix.com/covid19db

## Acknowledgements

We thank the employees of Helix, employees of Illumina, members of the CDC SPHERES consortium, and members of the Andersen Lab for discussion and help with logistics. We thank the healthcare workers, frontline workers, and patients who made the collection of this SARS-CoV-2 dataset possible and all those who made genomic data available for analysis via GISAID (**Supplemental Table S3**). This work has been funded by CDC BAA contracts 75D30121P10258 (Illumina, Helix) and 75D30120C09795 (GWY, RK, LCL, KGA), NIH NIAID 3U19AI135995-03S2 (MAS, KGA), U19AI135995 (MAS, KGA), U01AI151812 (KGA), NIH NCATS UL1TR002550 (KGA), the Innovative Genomics Institute (CYC), and the New Frontiers in Research Fund provided by the Canadian Institutes of Health Research (CYC).

## Methods

### Ethical statement

Helix data analyzed and presented here were obtained through IRB protocol WIRB#20203438, which grants a waiver of consent for a limited dataset for the purposes of public health under section 164.512(b) of the Privacy Rule (45 CFR § 164.512(b)). This work was also evaluated and approved by the Institutional Review Board at The Scripps Research Institute under IRB protocol IRB-15-6664. The work was conducted under a waiver of consent and received a non-human subjects research designation (category 4 exemption) because this research was performed with remnant clinical diagnostic specimens. All samples were de-identified before receipt by the study investigators.

### Helix COVID-19 Test Data

The Helix COVID-19 Test (EUA 201636) was run on specimens collected across the US, and results were obtained as part of our standard test processing workflow using specimens from anterior nares swabs. The Helix COVID-19 Test is based on the Thermo Fisher TaqPath™ COVID-19 Combo Kit, which targets three SARS-CoV-2 viral regions (N gene, S gene, and ORF1ab). Since samples are deidentified prior to analysis, and some individuals may test more than once, there may be some duplicate individuals in the analyses that follow that could cause deviation from the true population fraction. We attempted to de-duplicate the samples by removing those with identical age+sex+ethnicity+zipcode+clade, which only revealed three sets of two B.1.1.7 samples. Test results from positive cases, together with a limited amount of metadata (including sample collection date, state, and RT-qPCR Cq values for all gene targets), were used to build the research database used here. The data used in this paper is available in Supplemental Table S3. Ongoing summary level data are viewable at https://www.helix.com/covid19db.

### SGTF and B.1.1.7 quantitative analysis

Following conservative approaches from prior studies ((Bal et al., 2020; Kara Steel And, 2021), we filtered our dataset for positive samples with strong amplification of the N gene (Cq < 27). SGTF was annotated to samples with no S gene detected (Cq = Null). While this approach removed true positive results, and likely some SGTF samples, the variable behavior of the assay at longer cycle times warrants strict filtering for analysis. We applied the Cq(N gene) < 27 filter to all positive samples prior to analysis. Date of sample collection from each patient’s Test Requisition Form was used as the sample date for all analyses. Sequenced samples reflect those collected from December 17, 2020 through January 18, 2021.

Since we enriched for B.1.1.7 by selecting SGTF samples for sequencing, proportion of B.1.1.7 in total positive tests were inferred using,

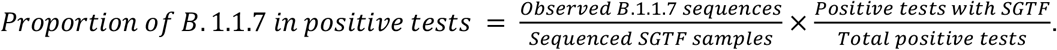

A logistic growth model was fit to the B.1.1.7 population prevalence over time using the *nls()* function in R. Following Voltz et al (Volz et al., 2021), increased transmissibility was estimated using a serial interval of 5 - 6.5 days using,

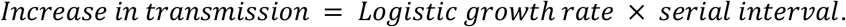

Since logistic growth rates are roughly exponential in the early phase, we estimate a rough constant doubling time using,

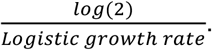

Data and code used for the analysis are available at https://github.com/andersen-lab/paper_2021_early-b117-usa.

### Helix/Illumina SARS-CoV-2 sequencing and consensus sequence generation

Residual samples remaining after reporting of the SARS-CoV-2 tests were selected for sequencing based on SGTF status and Cq(N gene) < 27 (note some early samples were selected up to Cq=35, though most were unable to produce usable sequence). Beginning in late December 2020, SGTF residual samples were saved for sequencing within 3 days of SARS-CoV-2 testing at Helix.

All samples were prepped and sequenced following the Illumina CovidSeq Test Instructions for Use (1000000128490 v01). RNA was extracted from 400 µl of patient sample using the Quick-DNA/RNA Viral MagBead kit (Zymo Research, # R2141) but was not treated with Proteinase K. During the Amplify cDNA step, the annealing temperature was reduced from 65°C to 63°C. Samples were sequenced using the NovaSeq 6000 Sequencing system S4 flow cell, which included the NovaSeq 6000 Sequencing System S4 Reagent Kit v1.5 (35 cycles) (Illumina, # 20044417) and the NovaSeq Xp 4-Lane Kit v1.5 (Illumina, # 20042337).

The NovaSeq flow cell output was further processed through the Illumina DRAGEN COVIDSeq Test Pipeline v1.3.0.28 to perform variant and consensus sequence generation for each sample. First flowcell output was demultiplexed into per-sample FASTQ sequences. Each sequence was then run through a DRAGEN kmer-based alignment algorithm. This algorithm utilized a kmer reference database to match kmers from the sequencing read to kmers from the SARS-CoV-2 reference genome (NCBI Accession NC_045512.2). To create the kmer reference list, the SARS-CoV-2 reference genome was split into 32 nucleotide (nt) kmers, and any kmers containing cross-reactivity were removed. To measure cross-reactivity, the kmer reference list incorporated the NCBI database of 100,000 genomes for human and animal pathogens in addition to the SARS-CoV-2 reference. Bat and pangolin viruses were excluded because of their similarity to the SARS-CoV-2 genome. Each of the reference kmers was labeled with a corresponding amplicon from either SARS-CoV-2 or external controls. If an amplicon contained at least 150 matches to SARS-CoV-2 reference kmers, the amplicon was considered detected. Variant calling and consensus sequence generation was performed for every sample with at least 90 SARS-CoV-2 virus amplicon targets.

Variant calling was performed by first aligning reads to the SARS-CoV-2 reference genome with the DRAGEN alignment module, then processing the aligned reads with the DRAGEN sort and duplicate removal modules and finally calling variants using the DRAGEN somatic “tumor-only” variant caller configured for haploid genomes. To generate a consensus sequence in FASTA format, detected sequence variants from the VCF output meeting the following criteria were applied to the SARS-CoV-2 reference sequence: a “PASS” entry in the FILTER column, the variant allele frequency ≥0.5, and the total filtered depth >10. Regions of sequence with coverage <10 were hard masked with N’s in the consensus sequence.

### Andersen lab at Scripps Research SARS-CoV-2 sequencing and consensus sequence generation

SARS-CoV-2 RNA was extracted from patient samples using either the MagMAX Viral/Pathogen II Nucleic Acid Isolation kit (Thermofisher, #A48383) or the Omega BioTek MagBind Viral DNA/RNA Kit (Omega Biotek, #M6246-03) according to manufacturers’ instructions. The extracted SARS-CoV-2 RNA was reverse transcribed using SuperScript IV VILO (ThermoFisher, #11756500). The virus cDNA was amplified in two multiplexed PCR reactions using ARTIC Network n-CoV-19 V3 primers and Q5 DNA High-Fidelity Master Mix (New England BioLabs, #M0492L) to generate tiled PCR amplicons. Libraries were then prepared for sequencing on Illumina platforms using Nextera XT (Illumina, #FC-131-1096). These libraries were sequenced on either Illumina NextSeq with a 500/550 Mid Output Kit v2.5 (Illumina, #20024908) or Illumina NovaSeq 6000 with an SP Reagent Kit v1.5 (Ilumina, #20028400) as 2×150 paired end reads (300 cycles). A subset of amplicon libraries were prepared for sequencing on the Nanopore MinION using KAPA HyperPrep kit (Roche, 07962363001) for end-repair and Ligation Sequencing Kit for adapter ligation (Oxford Nanopore, #SQK-LSK109). These libraries were individually sequenced on MinION R9 flow cells.

Consensus sequences from nanopore data were assembled using the arctic-ncov2019 pipeline (https://github.com/artic-network/artic-ncov2019). Consensus sequences from Illumina data were assembled using an inhouse Snakemake (Köster and Rahmann, 2012) pipeline with bwa-mem (Li, 2013) and iVar v1.2.2 (Grubaugh et al., 2019b).

### Chiu lab at UCSF SARS-CoV-2 sequencing and consensus sequence generation

Nasopharyngeal (NP) swab samples were prepared using 100 L of primary sample mixed with 100L DNA/RNA shield (Zymo Research, #R1100-250). The 1:1 sample mixture was then extracted using the Omega BioTek MagBind Viral DNA/RNA Kit (Omega Biotek, # M6246-03) on KingFisherTM Flex Purification System with a 96 deep well head (ThermoFisher, 5400630). Extracted RNA was reverse transcribed to complementary DNA and tiling multiplexed amplicon PCR was performed using SARS-CoV-2 primers according to a published protocol (Quick et al., 2017) and reagents from the Protoscript First Strand cDNA Synthesis Kit (New England Biolabs, #E6560L) and the Q5 Hot Start High-Fidelity DNA Polymerase Kit (New England Biolabs, #M0493L). Amplicons were ligated with adapters and incorporated with barcodes using NEBNext Ultra II DNA Library Prep Kit for Illumina (New England Biolabs, # E7645L). Libraries were barcoded using NEBNext Multiplex Oligos for Illumina (96 unique dual-index primer pairs) (New England Biolabs, # E6440L). Amplicon libraries were then sequenced on either Illumina MiSeq or Novaseq 6000 as 2×150 paired-end reads (300 cycles) with the MiSeq Reagent Kit V2 (Illumina, # MS-102-2002) or the NovaSeq 6000 Sequencing System S4 Reagent Kit v1.5 (Illumina, # 20028312), respectively.

Genome assembly of viral reads and variant calling were performed using an in-house automated bioinformatics pipeline as previously described (Deng et al., 2020).

### Phylogenetic analyses

Consensus FASTA for each sample was used as input to NextClade v0.12.0 (Hadfield et al., 2018) and Pangolin v2.0 (O’Toole et al., 2021) to label phylogenetic clades and lineages based on (Rambaut et al., 2020b), respectively. Samples identified as B.1.1.7 were selected for further phylogenetic analysis.

We downloaded all 26,064 sequences from the B.1.1.7 lineage and 21,032 sequences from the B.1.1 lineage, which forms the immediate outgroup of the B.1.1.7 lineage, from GISAID as of January 24, 2021. We combined this with our dataset of 212 B.1.1.7 sequences sampled in the U.S. and downsampled the combined dataset to 4,158 sequences based on collection date and country. (See **Supplemental Table S2** for GISAID acknowledgements.) We constructed a maximum-likelihood tree using this dataset under a HKY nucleotide substitution model implemented in iqtree2 (Minh et al., 2020). Using this phylogeny, we selected 504 sequences to retain, including all the sequences in our dataset (n=212) and a selection of global sequences to represent global diversity of the B.1.1.7 lineage (n=291 and NC_045512 as an outgroup root). We estimated the time-resolved phylogeny using a HKY nucleotide substitution model with discrete-gamma distributed rate variation under an uncorrelated relaxed clock model implemented in BEASTv1.10.5pre (Suchard et al., 2018). We used a relatively uninformative conditional reference prior on the overall clock rate (Ferreira and Suchard, 2008), an exponentially growing population prior over the unknown tree and the BEAGLE library to improve computational performance (Ayres et al., 2019). We let the analysis run for 100 million steps and discarded the first 10 million as burn-in. The effective sample size of all scientifically relevant parameters was above 200. The phylogenetic tree was visualized using baltic (https://github.com/evogytis/baltic). The BEAST XML and code to process the analysis are available at https://github.com/andersen-lab/paper_2021_early-b117-usa.

## Supplemental Information

**Figure S1.**
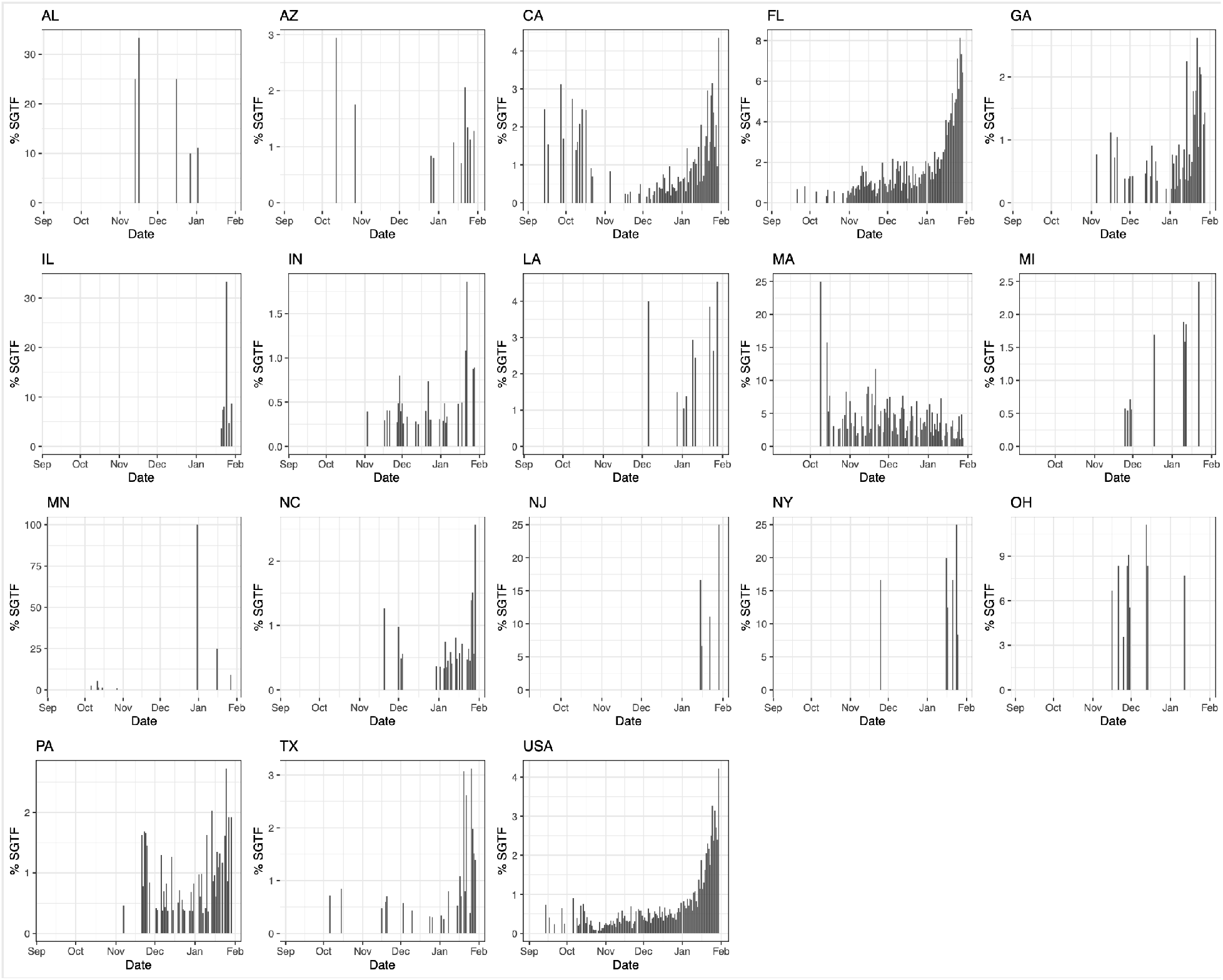
Percentage of SGTF for states with >500 SARS-CoV-2 positive tests at Helix since October 2020. Proportion of SGTF in daily positive tests with Cq(N gene) < 27, in states with >500 positive tests from October 15, 2020 to January 30, 2021. Given the average nationwide %SGTF of 0.6% during the studied period, rolling averages for states with fewer than 500 positive samples during the total sampled time period may be unreliable.

**Figure S2.**
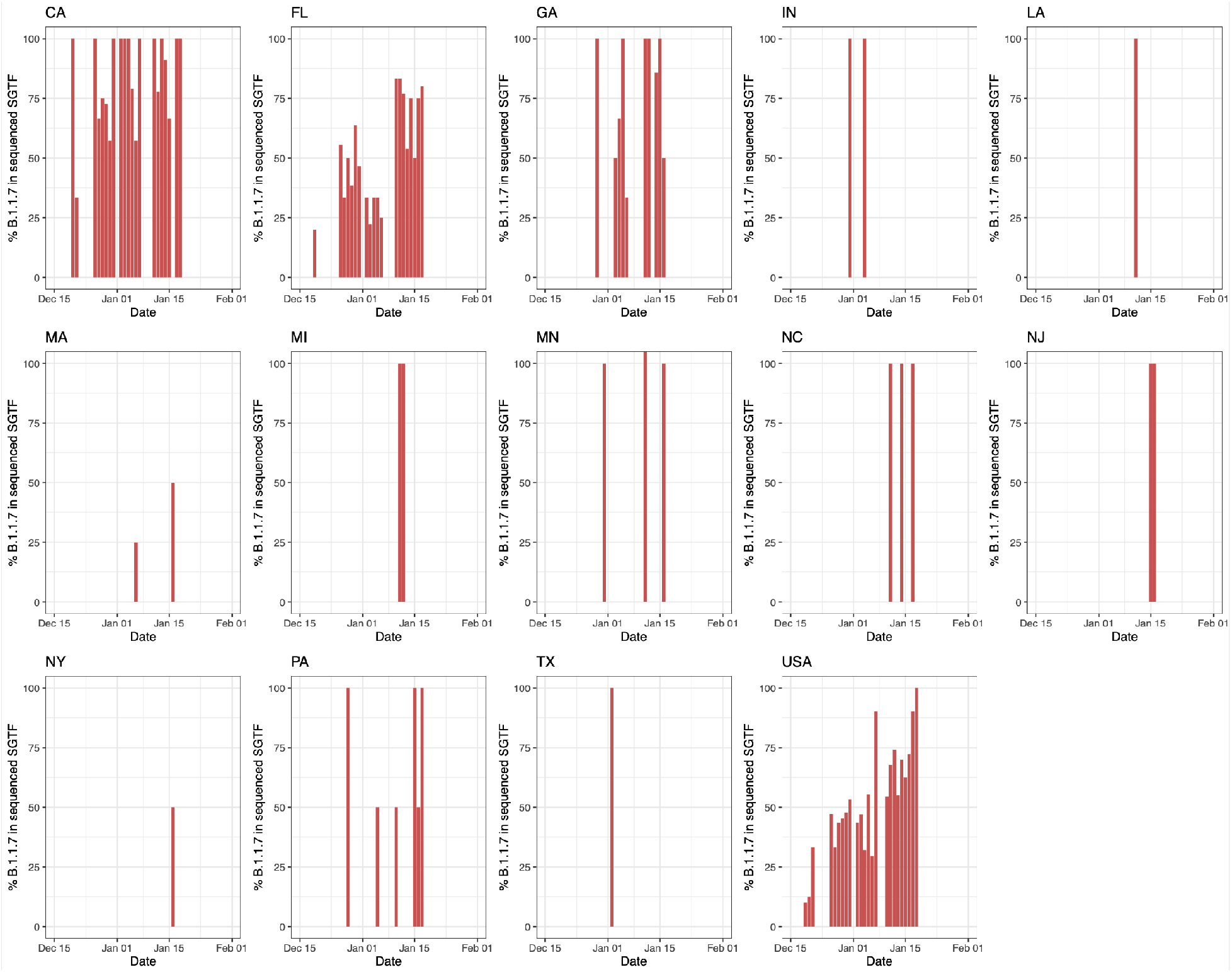
Percentage of B.1.1.7 in sequenced samples with SGTF over time in states with >500 SARS-CoV-2 positive tests at Helix since October 2020 and across the United States (USA).

**Table S1. Sequence ids, lineage, state, and collection date for samples analyzed in this study**.

**Table S2. GISAID acknowledgment table**

**Table S3. Raw Data on test results, SGTF, and B117 status grouped by State of residence and collection date, used in Figure 1**. We additionally provide number of sequences and number of sequenced SGTFs per date per state.

