## Supplementary figures and images for "Genomic epidemiology identifies emergence and rapid transmission of SARS-CoV-2 B.1.1.7 in the United States"

### Supplemental Figure S1 - Percentage of SGTF for states with >500 positive tests

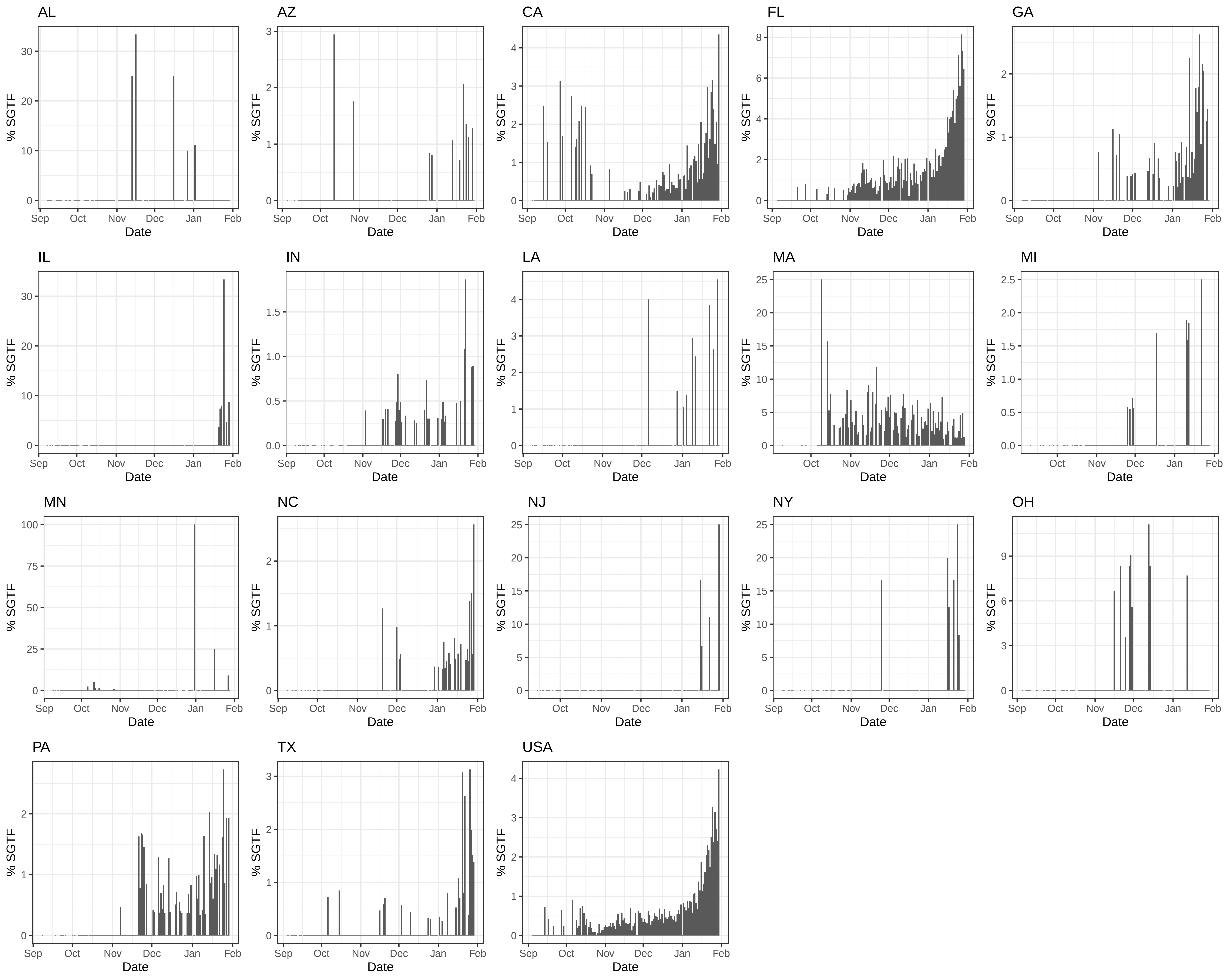

### Supplemental Figure S2 - Percentage of B.1.1.7 in sequenced samples with SGTF over time

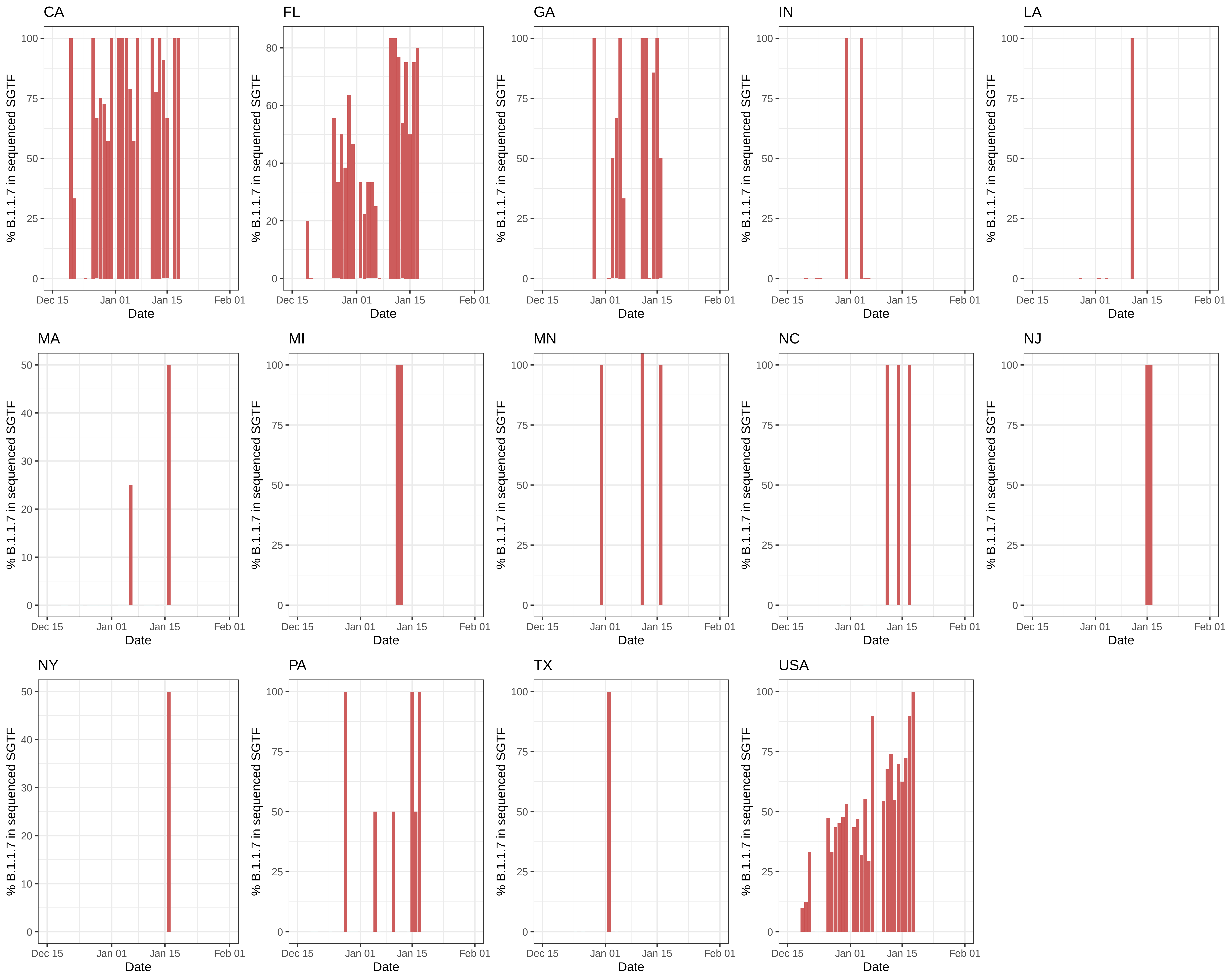
